# Analysis of sex disparities in under-five mortality rates in Ghana: Insights from vector autoregressive modeling

**DOI:** 10.1101/2023.02.17.23286087

**Authors:** Nana Owusu Essel, Simon Kojo Appiah, Isaac Adjei Mensah

**Author notes:** Corresponding author: Simon Kojo Appiah, PhD, Department of Statistics and Actuarial Science, College of Science, Kwame Nkrumah University of Science and Technology, Private Mail Bag, University Post Office, Kumasi, Ghana.

## Abstract

International monitoring organizations call for child mortality indicators to be disaggregated by gender. However, there remains a paucity of studies, especially, from the sub-Saharan region aimed at producing accurate forecasts of child mortality indicators with their sex variations. This study aims at investigating disparities in indicators of childhood mortality rates by sex in Ghana by employing vector autoregressive (VAR) model to analyze jointly annual recorded data on total, male and female under-five mortality rates (TU5MR, MU5MR, FU5MR, respectively). The results show gradual declining under-five mortality trends among sexes in both the historical and forecasted rates. The trivariate traditional and instantaneous Granger causality analyses found that any of the mortality indicators Granger causes the other two combinations, except TU5MR to MU5MR and FU5MR. The forecast error variance decomposition analyses revealed that FU5MR was the most exogenous variable while long-term impulse response function analyses indicated that unit shocks in FU5MR significantly increased TU5MR. The VAR(2) model forecast constructed revealed that contrary to recent predictions based on wider interval data derived from demographic health surveys, Ghana may meet the SDG 3.2.2 if ongoing efforts are sustained and that focusing policies and interventions on reducing FU5MR would largely contribute to reducing TU5MR in Ghana.

**Ethical considerations:** Not applicable. This study did not require ethics approval or consent for participation.

## 1. Introduction

Sex remains a primary factor for disaggregating indicators of childhood mortality for analytic and monitoring purposes [1]. Most countries worldwide have seen substantial reductions in childhood mortality rates in recent times. Despite this, it remains crucial to ensure that these survival improvements are equitable to children as much as possible. Up to the age of 5 years, survival tends to favor girls over boys, with a mortality sex ratio greater than 1 [2]. Due to changes in the distributions of associated causes of death, generally advantageous to girls at lower mortality rates, their survival tends to increase as total mortality rates decline. After early infancy, however, girls do not show the same advantages in survival due to infectious diseases; thus, sex differentials in child mortality tend to be lower than those of infant mortality, with those of under-five mortality taking values between the two [3]. Excess female mortality could indicate the presence of factors outweighing the expected biological survivorship conferred on girls during these ages, as would occur if girls are subjected to unequal access to healthcare, nutrition, or sex-related biases because of community preference for males [4]. On the other hand, higher-than-expected sex mortality ratios, indicative of inequities against male children, might point to a change in mortality to an extent greater than biologically expected. For instance, Drevenstedt et al. [5] identified that a declining prevalence of infectious diseases and a concurrent rise in perinatal causes led to an increase in sex ratios of infant mortality, following which improvements in obstetric and perinatal care resulted in its decline.

Differences in sex mortality ratio have been attributed to an interplay among biological, social, and environmental factors [5]. Despite contributing to sex disparities in adult mortality indicators, lifestyle and behavioral factors are improbable determinants of infant mortality [5]. Further, other determinants of typically high or low mortality sex ratios may exist, including disparities in the treatment of girls compared to boys as reported in studies conducted in Asian countries [6,7]. Identifying countries with similarly unusual levels of sex disparities in childhood mortality, suggesting possible differences in treatment, is key to monitoring sex discrimination [2]. Also, although the sex mortality ratio is generally considered an important indicator of equity in the healthcare of children, its effective use has been restricted by the lack of clear-cut ideal ratios [8]. One of the earliest applications of vector autoregressive (VAR) modeling to child mortality investigated the dynamic relationship between infant mortality and fertility, and their connection with demographic transition [9]. More recently, VAR models have been applied in econometrics to investigate the impact of healthcare spending on infant and child mortality [10] and in public health to identify the roles played by socioeconomic well-being and female sex on childhood mortality in Bangladesh [11]. Rajab et al. [12] examined the spread of COVID-19 infection in the UAE, Saudi Arabia, and Kuwait using a VAR model to jointly forecast the number of new cases and deaths while Li and Lu [13] proposed a spatio-temporal version of VAR (STAR) to forecast mortality rates according to age, depending on historical and neighboring values.

To the best of our knowledge, no previous study has focused on trends in long-term sex disparities in childhood mortality indicators in Ghana or has aimed at forecasting these disparities. Understandably, such a task aimed at estimating trends in sex disparities, especially in a setting such as Ghana with unreliable vital registration services, cannot be expected to be straightforward, and these data are either limited or noisy compared to total estimates without regard to sex. The present article, therefore, aims to fill the existing gap in the literature by utilizing data from the UNICEF Data Warehouse [14] from 1970 to 2020 to examine trends in sex differentials in under-five mortality rate (U5MR) using a VAR model and perform a 5-year forecast of each indicator to inform the effectiveness of previous and existing policies directed toward reducing inequalities in childhood mortality. VAR is typically used to model the joint dynamic behavior of multivariate time series of events such as deaths or infectious diseases. The VAR model generalizes the univariate autoregressive model for predicting time series of U5MRs as a vector, considering each component or variable as a linear function of historical lags in its series and those of other variables. The advantage of the VAR model is that it allowed us to incorporate total and sex-wise mortality rates in a single model, thereby yielding a much better paradigm for prediction [12]. In this case, the time series of total, male, and female U5MRs (TU5MR, MU5MR, and FU5MR, respectively) are assumed to be related and combined to obtain a joint forecasting model for more accurate forecasts, taking into account the cross-correlation between the mortality rate datasets.

The remaining aspects of the paper are structured into four sections. Section 2 of this paper focuses on constructing an adequate VAR model for TU5MR, MU5MR, and FU5MR as system variables. After constructing and diagnosing such a model, Section 3 presents our approach to making statistical inferences about the mortality model by way of impulse response function (IRF) analysis, Granger causality testing, cointegration analysis, and performing a 5-year forecast using the VAR process. Section 4 gives a detailed discussion on the study findings, and also presents the strengths and limitations of the study while Section 5 finalizes the paper by drawing conclusions from the study.

## 2. Materials and methods

### 2.1. Data source and variable descriptions

We used openly-available annual U5MR data on Ghana, spanning the period 1970–2020, obtained from the UNICEF Data Warehouse (https://data.unicef.org/dv_index/), last accessed in August 2022. The warehouse is a leading source of data on the pediatric population and is involved in the maintenance of many internationally valid indicators of healthcare in this population. Datasets contained in this warehouse are readily available for almost all countries, with data mostly spanning several decades. Here, in line with the WHO [15] definition, TU5MR is defined as the probability (calculated in rate form, per 1000 live births) that a child born between 1970 and 2020 would die from any cause before attaining the age of 5 years. The sex-wise composites FU5MR and MU5MR are the estimated probabilities (per 1000 live births) that a girl or boy born in Ghana between 1970 and 2020 would die before their fifth birthday. Since one goal of the study was to provide five-year forecasts of three U5MRs (TU5MR, MU5MR, and FU5MR), time plots of these datasets are shown in Fig. 1. These plots demonstrated a noticeable trend in the data, indicative of non-stationarity in the three mortality indicators under study.

**Fig. 1.**
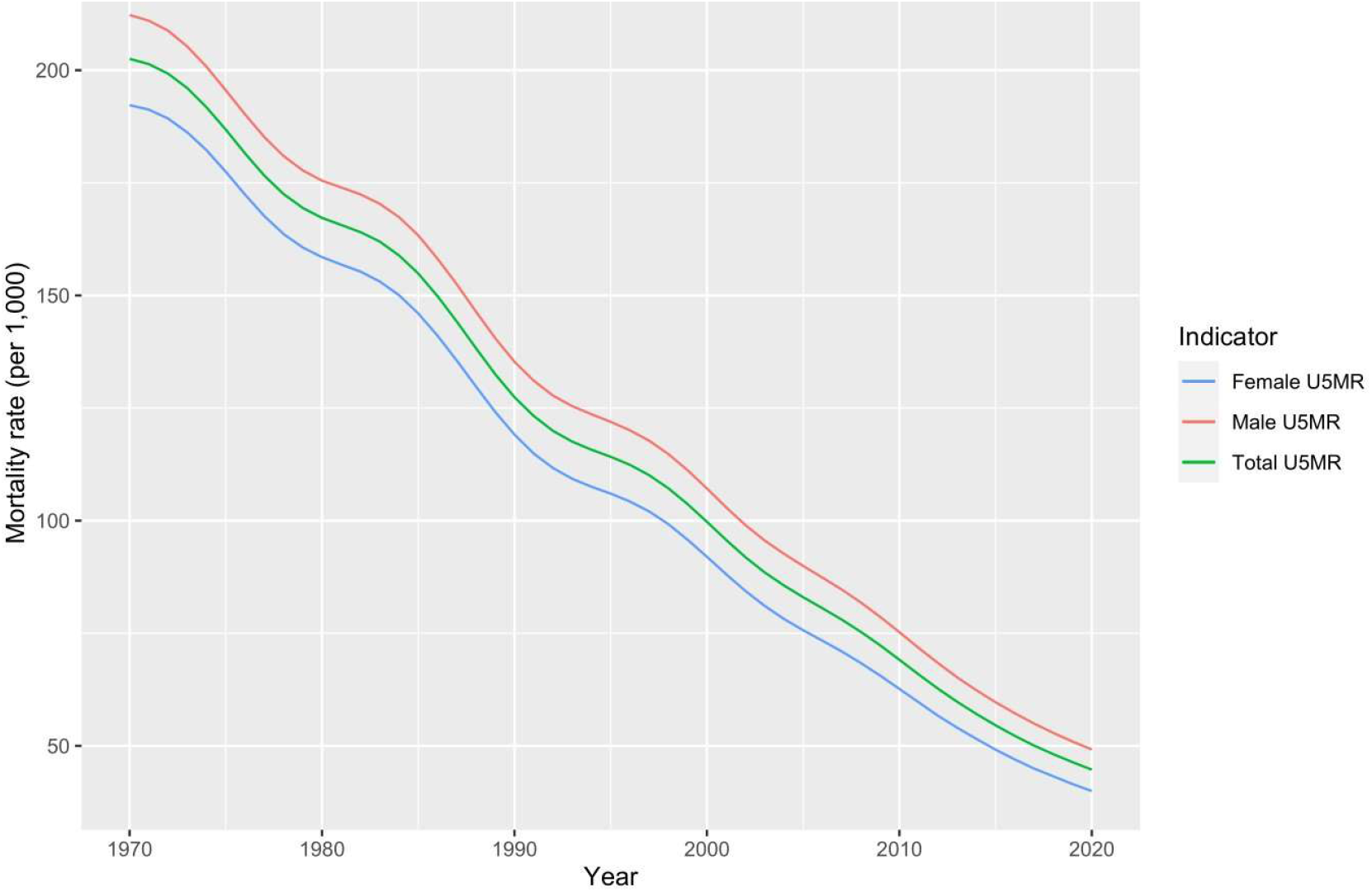
Time series plots showing historical variations in total, male, and female under-five mortality rates (U5MRs) in Ghana, 1970–2020

All statistical analyses were performed using RStudio 2022.07.2 in the R 4.2.2 environment (The R Development Core Team, Vienna, Austria) with the *forecast, urca, vars, mFilter, tidyverse*, and *tseries* packages. Statistical tests with *p*-value <0.05 were considered alongside their 95% confidence intervals (CIs) to indicate statistical significance.

### 2.2. Vector autoregressive modelling

The VAR model is one of the most successfully used and flexible approaches to multivariate time series analysis. It is a natural extension of the univariate autoregressive model, describing how a set of *k* (endogenous) variables, in this case, the three under-five mortality time series (TU5MR, MU5MR, and FU5MR), evolve over time. The main steps involved in a typical VAR modeling as employed for the data analysis in this study are schematically presented in Fig. 2. The detailed methodology of VAR analysis is presented in the following sequel.

**Fig. 2.**
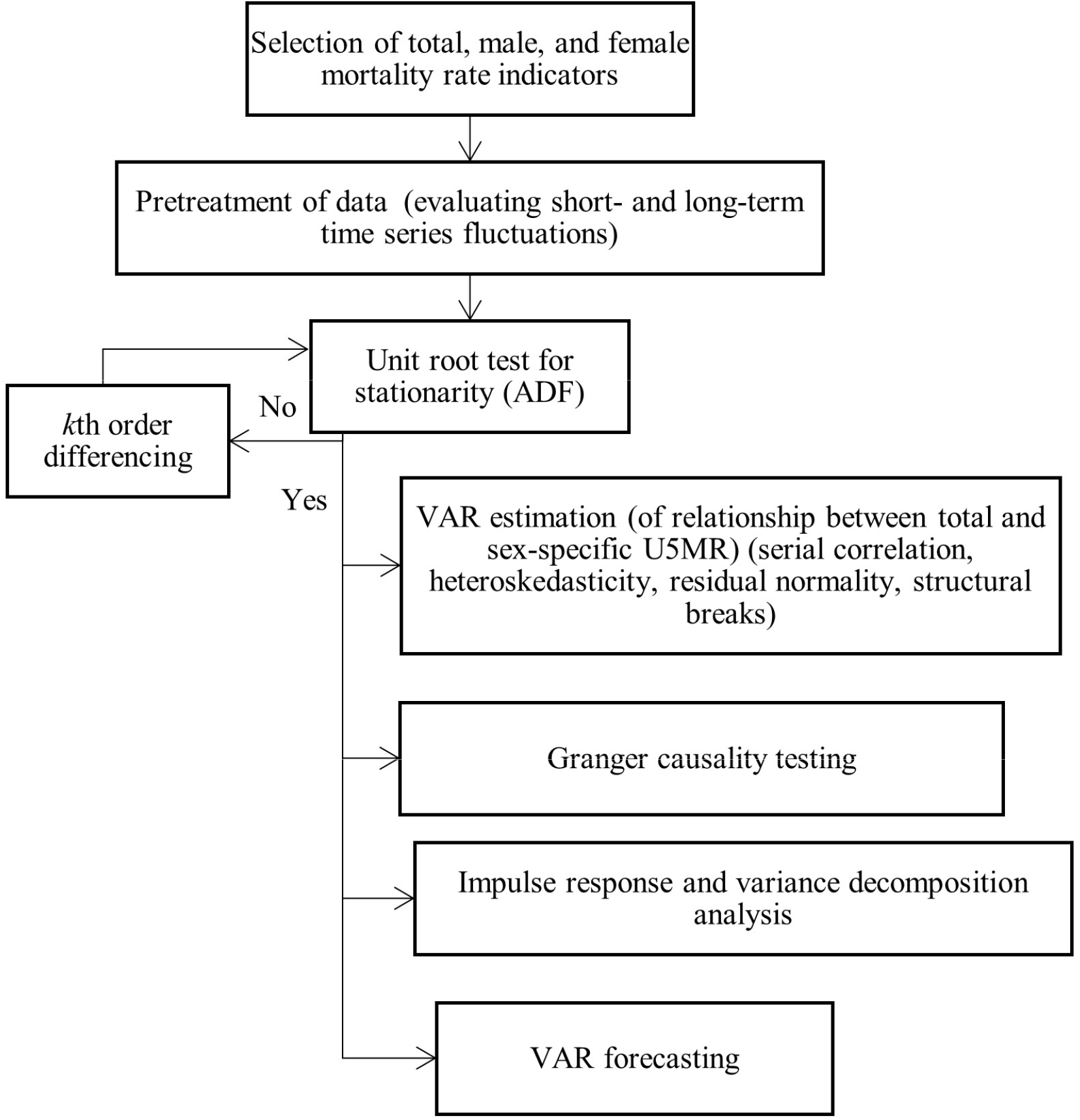
Basic flow chart showing the steps in VAR modeling of total, male, and female U5MR VAR, vector autoregression; U5MR, under-five mortality rate; ADF, Augmented Dickey–Fuller

#### 2.2.1. VAR model specification and estimation

The structure of a VAR model is that each variable is a linear function of past lags of itself and other variables. Hence, for a given set of *k* variables and time period *t* = 1, …, *T*, we have the vector *Y*_*t*_ = (*y*_l*t*_, *y*_2*t*_, …, *y*_*kt*_)^*T*^, where *Y*_*i,t*_ is the *i*th variable at time *t*. A VAR model is described by the number of preceding time periods the model utilizes, called *order*. A VAR model of order *p, VAR*(*p*), is defined by:

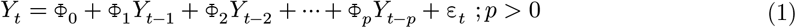

where *Y*_*t*_ is a *k* × 1 vector of the dependent variables; Φ_O_ is a *k* × 1 vector of constants; Φ_*i*_; *i* = 1, 2, …, *p* are *k* × *k* coefficient matrices; and ε_*t*_ is *k* × 1 serially uncorrelated error vector with multivariate normal distribution mean vector 0 and a nonsingular covariance matrix Σ. The *VAR*(*p*) model parameters Φ = (Φ_O_, Φ_l_, Φ_2_, …, Φ_*p*_) and Σ are estimated by the ordinary least squares (OLS) procedure following [16–18]:

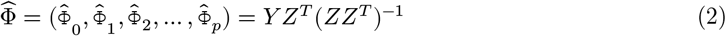

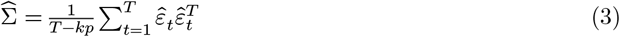

where *Y* = (*Y*_l_, *Y*_2_, …, *Y*_*T*_)^T^, *Z* = (*Z*_O_, *Z*_l_, *Z*_2_, …, *Z*_*T−l*_), *Z*_*T−l*_ = (*Y*_*t*−1_, *Y*_*t*−2_, …, *Y*_*t−p*_)^*T*^, 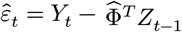, and 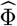 is consistent and asymptotically distributed as*N*_*k*_(0, Σ). The order *p* is determined by fitting *VAR*(*p*) models with orders *p* = 0, 1, …, *p*_*max*_ and then choosing the value of *p* to minimize various information criteria including the Akaike information criterion (AIC), Schwarz-Bayesian information criterion (BIC), and Hannan–Quinn criterion (HQ), defined by:

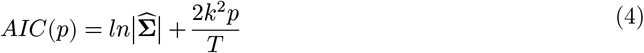

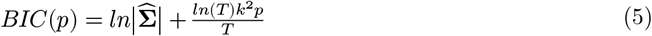

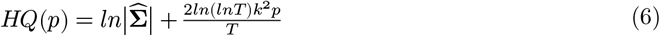

#### 2.2.2. Unit root and cointegration tests

VAR model validity depends on the stationarity of its composite time series [19]. For example, regressing nonstationary TU5MR on MU5MR and FU5MR would yield a so-called spurious regression model in which ordinary least-squares (OLS) estimates and *t*-statistics would indicate the existence of a relationship, when in reality, this is not the case; thus, the results obtained via such an approach would be erroneous. The unit root tests, specifically, the Augmented Dickey– Fuller (ADF) and Phillips–Peron (PP) tests, are employed to establish stationarity. The ADF statistic is used to investigate the null hypothesis that a unit root is present in the time series against the alternative that no unit roots are present, and is usually applied to evaluate stationarity or trend stationarity. The test itself is an augmented version of the traditional Dickey–Fuller test applied to a larger and more complex set of time series. The ADF test statistic returns a negative number, with more negative values providing stronger evidence for the rejection of the null hypothesis at a predefined confidence level. The ADF test is executed via a similar approach as that used in the Dickey–Fuller test but is applied to the model:

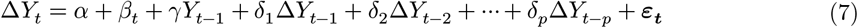

where *α* is a constant, *β* is the coefficient on a time trend *t, p* is the lag order of the autoregressive process, *γ* is the coefficient of the response variable with lag order 1, *δ*_1_, *δ*_2_, …, *δ*_*p*_ denote the parameter estimates with respect to the differenced response variables with lag-orders *t* − 1, *t* − 2, …, *t* − *p*, and ∆ is a difference operator. By including lags of order *p*, the ADF test permits application to higher-order autoregressive processes; thus, the lag length *p* needs to be set. One approach is to test down from high orders and examine *t*-values on coefficients while another approach involves examining the AIC and BIC values. The unit root test is then performed under the null hypothesis of *γ* = 0 against the alternative that *γ* > 0. Upon calculating the test-statistic 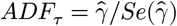, we compare it to the relevant critical values of the Dickey–Fuller test, and reject the null hypothesis if *ADF*_*τ*_ is more negative than the critical value.

The PP test can also be used to investigate the null hypothesis that a time series is integrated of order 1. It is structured on the Dickey–Fuller test of the null hypothesis *ρ* = 1 based on the model:

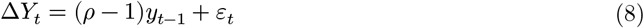

with ∆ representing the first difference operator. Similar to the ADF test, the PP test addresses the issue that the order of autocorrelation for the process generating data for *Y*_*t*_ might be higher than admitted in the test equation, rendering *Y*_*t*−l_ endogenous and thereby invalidating the Dickey– Fuller *t*-test. Unlike the ADF test which handles this issue by introducing lags of ∆*Y*_*t*_ as regressors in its equation, the PP test corrects the *t*-test statistic non-parametrically. It is robust with respect to unspecified autocorrelation and heteroscedasticity in the disturbance process of the test equation [18].

Specifically, the three mortality indicators (variables) employed would be considered cointegrated if any of their linear combinations is stationary. Cointegration in such a time series vector would be assessed on the intuition that they have a long-term equilibrium relation [20–22]. To identify the presence of cointegration, and thus, the number of cointegrated time series, we applied the Johansen approach using the relevant trace and maximum eigenvalue statistics. In applying the Johansen approach, we consider a *VAR*(*p*) model with integration order 1, *I*(1), which is reformulated as:

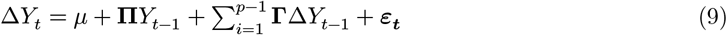

with 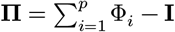 and 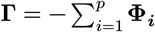,. If the coefficient matrix ∏ reduces to *r* < *k*, there must exist matrices ***α*** and ***β*** with dimensions *k* × *r* and rank *r* each such that **П** = ***αβ*** is stationary. Assuming that *r* is the number of cointegration relationships, *α* is the number of adjustment parameters in the vector error correction model, and each column of ***β*** is a cointegrating vector, we can infer that for a given *r*, the maximum likelihood (ML) estimator of ***β*** defines the combination of *Y*_*t*−l_ yielding the *r* largest canonical correlations of ∆*Y*_*t*_ with *Y*_t−l_ upon correcting for lagged differences and deterministic variables when present. Johansen [23] proposed two forms of likelihood ratio tests for the significance of these canonical correlations involving the reduction of the ranks of **П**. The trace and maximum eigenvalue test statistics are given in Equations Error! Reference source not found. and Error! Reference source not found. below:

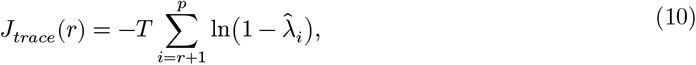

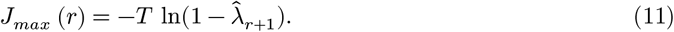

where *T* is the sample size and 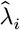 is the *i*th largest canonical correlation.

*J*_*trace*_(*r*) tests the null hypothesis of *r* cointegration vectors versus the alternative of *k* cointegrating vectors, whiles *J*_*max*_(*r*) tests the null hypothesis of *r* cointegrating vectors versus the alternative of *r* + 1 cointegrating vectors. Although Johansen’s methodology is usually applied to systems characterized solely by *I*(1), theoretically, the presence of stationary variables is not an issue and there is little need for pre-testing to establish the order of integration [23].

#### 2.2.3. Structural analysis

The *VAR*(*p*) model has several parameters which can be difficult to interpret due to complex interactions and feedbacks between the variables. A standard VAR modeling often reports on structural analysis, which involves Granger causality tests, IRFs, and forecast error variance decompositions (FEVDs).

##### 2.2.3.1. Granger causality

The Granger causality tests are performed to study the causal relationship among the variables in the system. It begins by comparing the following unrestricted models:

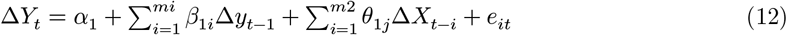

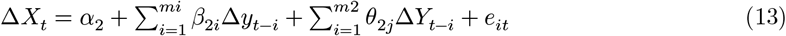

with the restricted models:

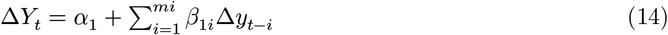

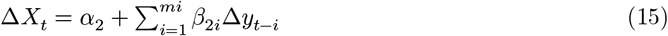

where, ∆*Y*_*t*_ and ∆*X*_*t*_ first-order forward differences of the variables; *α, β*, and *θ* are the parameters we need to estimate; *e*_*it*_ represents standard random errors, and *m* is the optimal lag order chosen using the information criteria. Equations Error! Reference source not found. are convenient for analyzing linear causal relationships among the variables. If *θ*_l_ is statistically significant and *θ*_2_ is not, we can say that changes in variable *Y* Granger cause changes in variable *X* or vice versa. If both *θ*_l_ and *θ*_2_ are statistically significant, *X* and *Y* have a bivariate causal relationship. If neither is statistically significant, neither changes in *Y* nor *X* have any effect on the other variable.

##### 2.2.3.2. Impulse response functions

The *VAR*(*p*) model also has the following Wold’s decomposition (or representation):

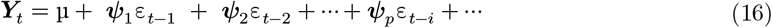

where **ψ**_*s*_ are *k* × *k* matrices with an (*i, j*)th element, 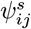, interpreted as the dynamic multiplier or impulse response:

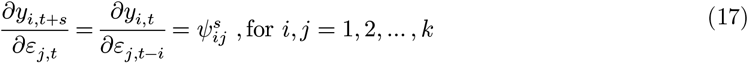

under the condition that *Var*(*ε*_*t*_) = **Σ** is equal to a diagonal matrix, implying that the elements of **ε**_*t*_ are uncorrelated [18]. To make the errors uncorrelated we estimate the triangular structural *VAR*(*p*) model:

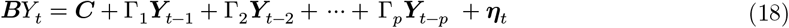

where ***B*** is a lower triangular matrix with 1s as diagonal elements and uncorrelated (or orthogonal) errors ***η***_*t*_, known as *structural errors*. Then, Wold’s decomposition of ***Y***_*t*_ based on orthogonal errors ***η***_*t*_ can be given by:

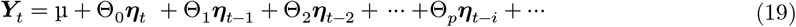

where Θ_O_ = ***B***^−l^ is the lower triangular matrix. The impulse responses to the orthogonal shocks ***η***_*t*_ are given by

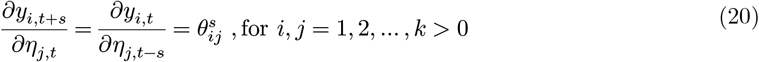

in which 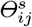 represents the (*i, j*)th element of Θ_*s*_. A plot of 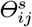 against *s* is referred to as the orthogonal IRF of *Y*_*i*_ with respect to ***η***_*t*_.

##### 2.2.3.3. Forecast error variance decompositions

The FEVD accounts for the portion of the variance of the forecast error in predicting ***Y***_*t,T+h*_ due to the structural shock *η*_*j*_. Using the orthogonal shocks ***η***_*t*_ the *h*-step ahead forecast error vector, with known VAR coefficients, may be expressed as

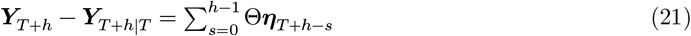

where for a particular *y*_*i,T+h*_, the forecast error has the form:

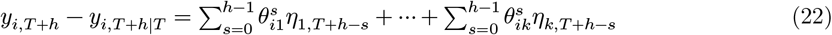

with variance due to orthogonality of the structural errors:

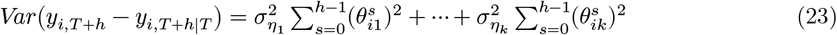

where 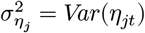 and the portion of the variance due to shock *η*_*j*_ is defined by:

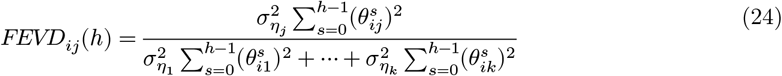

called the FEVD, which depends on recursive causal ordering used to identify structural shocks *η*_*j*_.

#### 2.2.3.4. VAR forecasting

After ensuring adequacy of the fitted *VAR*(*p*) models, the *h*-step ahead forecasts are made using the best linear form *Y*_T +h_ given by:

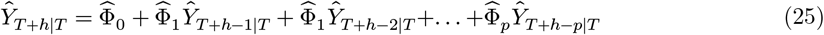

with forecast error:

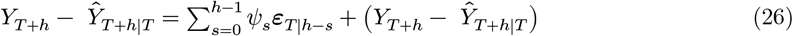

and forecast covariance matrix:

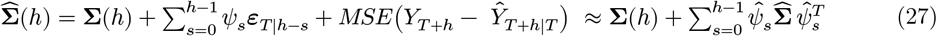

where (*Y*_*T+h*_ − *Ŷ*_*T +h|T*_) is the error due to estimating *VAR*(*p*) parameters and 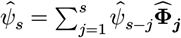. The (1 − *α*)100% CIs for the components of *Y*_*T+h*_ are computed by:

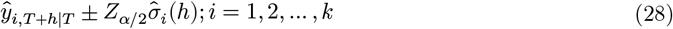

where 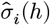 is the square root of the *i*th diagonal element of 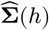.

### 2.3. VAR model diagnostics

#### 2.3.1. Serial correlation analysis

The Portmanteau test was performed to check absence of autocorrelation in the residuals of the estimated VAR(2) model generated for U5MRs. The null hypothesis is tested for residual no serial correlations (autocovariances are zero) with test-statistic:

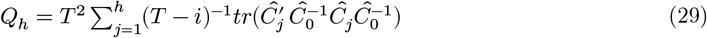

where 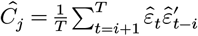, and has approximately 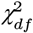 distribution with degrees of freedom *df* = *hk*^2^ [16,24]

#### 2.3.2. Model heteroskedasticity

The presence of autoregressive conditional heteroskedasticity (ARCH) tends to invalidate VAR parameter estimates of *VAR*(*p*) and hence undermine their efficiency and inferences performed on them. To test for ARCH effects, a multivariate Lagrange multiplier (MLM) test based on the auxiliary regression model was applied:

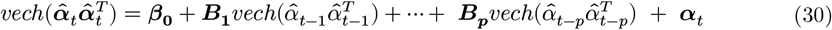

where ***β***_0_ is a 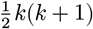-dimensional vector, ***B***_l_, ***B***_2_, …, ***B***_*p*_ are 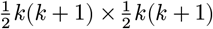 matrices, and ***α***_*t*_ is a spherical error vector process. The hypothesis test, *H*_O_: ***B***_l_ = ***B***_2_ = … = ***B***_*p*_ = **0** (lack of ARCH effects in residuals) versus *H*_l_: ***B***_*j*_ ≠ **0**; for *j* ≥ 1 is performed with test-statistic:

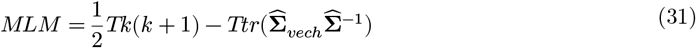

where 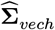 is the estimator for the error covariance matrix in model (30) while 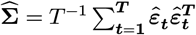is the estimator of the error covariance matrix in model (1), which is asymptotically distributed as 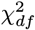 with degrees of freedom 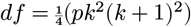.

#### 2.3.3. Tests for normality and structural stability in VAR(*p*) residuals

Although VAR model residuals and observed variables do not need to be normally distributed for the model to be valid, normality in residual distribution is of interest as it facilitates predictive inference. Multivariate versions of the Jarque–Bera, skewness, and kurtosis tests were implemented on the VAR(2) model residuals standardized via Cholesky decomposition of the covariance matrix of centered residuals. Structural breaks in the residuals of the under-five mortality rates were tested for the fitted VAR(2) model based on the first *K* observations for *k* = *k*_l_, *k*_2_, …, *k*_*K*_, where *k*_l_ is characterized by the necessary degrees of freedom required for estimation. Plots of each recursive estimate with respective standard errors or confidence limits for *k* = *k*_l_, *k*_2_, …, *k*_*K*_ can provide useful information on the possibility of structural breaks. Subsequently, an OLS cumulative sum (OLS-CUSUM) approach was used to plot empirical fluctuation processes to visualize structural variations in all three mortality indicators. The OLS-CUSUM test statistic could alternatively have been used, typically computed as:

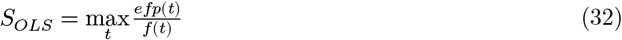

in which *efp*(*t*) represents the empirical fluctuation process and *f*(*t*) is constrained on the boundary *b*(*t*) = *λ*f(*t*). We would then entertain the null hypothesis if |*S*| < |*b*(*t*)| [25].

## 3. Results

### 3.1. Stationarity and cointegration tests

The unit root tests presented in Section 2.2 were applied to determine whether the three nonstationary mortality time series require differencing to render them stationary or regression on deterministic functions of time. The ADF and PP tests were performed for each U5MR variable under the null hypothesis of non-stationarity at a 5% significance level, the results of which are presented in Table 1. The results confirmed that all three variables had nonstationary levels (*p-* values ≥0.05). We, thus, incorporated trends into the model generated, and needed not detrend each series as the *vars* library is capable of fitting *VAR*(*p*) models with trend as a deterministic regressor. Applying the *VARselect* function in the *vars* package showed that 2 lags were optimum based on the selection criteria, AIC, BIC, and HQ, as outlined in equations 4–6. The Johansen cointegration test was further performed to examine whether there exist long-run relationships (cointegration) among the employed time series mortality models. Table 2 outlines the critical and test statistic values derived from the Johansen cointegration procedure. From the likelihood ratio (trace) and maximum eigenvalue tests of cointegration, we adjudged that at most two of the U5MR time series were cointegrated at ranks *r* ≤ 1 and *r* ≤ 2, and significance levels 10%, 5%, and 1%.

**Table 1.** Results of Augmented Dickey–Fuller (ADF) and Phillips–Perron unit root tests

| Time series variables | ADF test |  |  | Phillips–Perron test |  |  |
| --- | --- | --- | --- | --- | --- | --- |
| | Test statistic | Lag order | $p$-value | $Z$ statistic | Truncation lag parameter | $p$-value |
| TU5MR | 1.2292 | 3 | 0.99 | -3.9248 | 3 | 0.8869 |
| MU5MR | 1.5760 | 3 | 0.99 | -5.1623 | 3 | 0.8112 |
| FU5MR | 1.6803 | 3 | 0.99 | -2.8169 | 3 | 0.9402 |

TU5MR, total under-five mortality rate; MU5MR, male under-five mortality rate; FU5MR, female under-five mortality rate

**Table 2.**
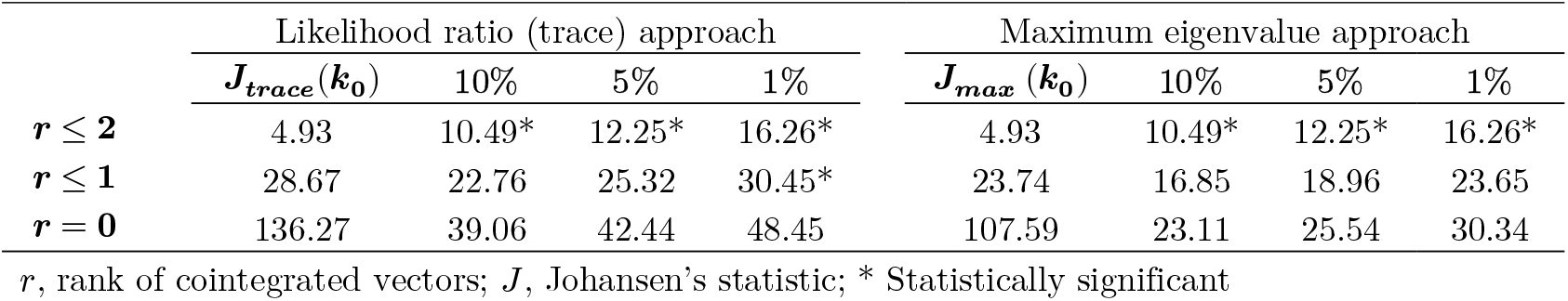
Results of Johansen cointegration test

|  | Likelihood ratio (trace) approach |  |  |  | Maximum eigenvalue approach |  |  |  |
| --- | --- | --- | --- | --- | --- | --- | --- | --- |
| | $J_{trace}(k_0)$ | 10% | 5% | 1% | $J_{max}(k_0)$ | 10% | 5% | 1% |
| $r \leq 2$ | 4.93 | 10.49* | 12.25* | 16.26* | 4.93 | 10.49* | 12.25* | 16.26* |
| $r \leq 1$ | 28.67 | 22.76 | 25.32 | 30.45* | 23.74 | 16.85 | 18.96 | 23.65 |
| $r = 0$ | 136.27 | 39.06 | 42.44 | 48.45 | 107.59 | 23.11 | 25.54 | 30.34 |

$r$ , rank of cointegrated vectors; $J$ , Johansen’s statistic; \* Statistically significant

### 3.2. VAR model estimation

#### 3.2.1. VAR model construction

Fitting the VAR(*p*) model Error! Reference source not found. by incorporating both constant and trend deterministic regressors, and without exogenous variables to the U5MRs data, we obtained the multivariate model estimates of TU5MR and its sex composites (MU5MR, FU5MR) via equations (2)–(6) with *p* = 2. The results parameters and their statistical significance are shown in Table 3. The results show that both the constant and trend deterministic regressors are statistically significant in all the three U5MR series (with *p*-values <2×10^−16^). Also, neither the first nor second lags of MU5MR, FU5MR, and TU5MR showed any significant influence on TU5MR and MU5MR in the overall model. On the other hand, the first lags of FU5MR significantly and nearly doubled subsequent trends in FU5MR in the established VAR(2) model (showing *β* = 1.771, *p*-value = 0.0189), in addition to significant effects of the constant and trend deterministic regressors.

**Table 3.**
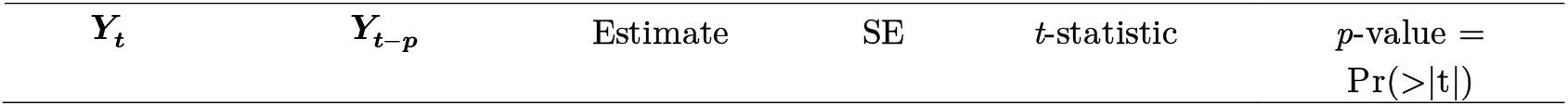
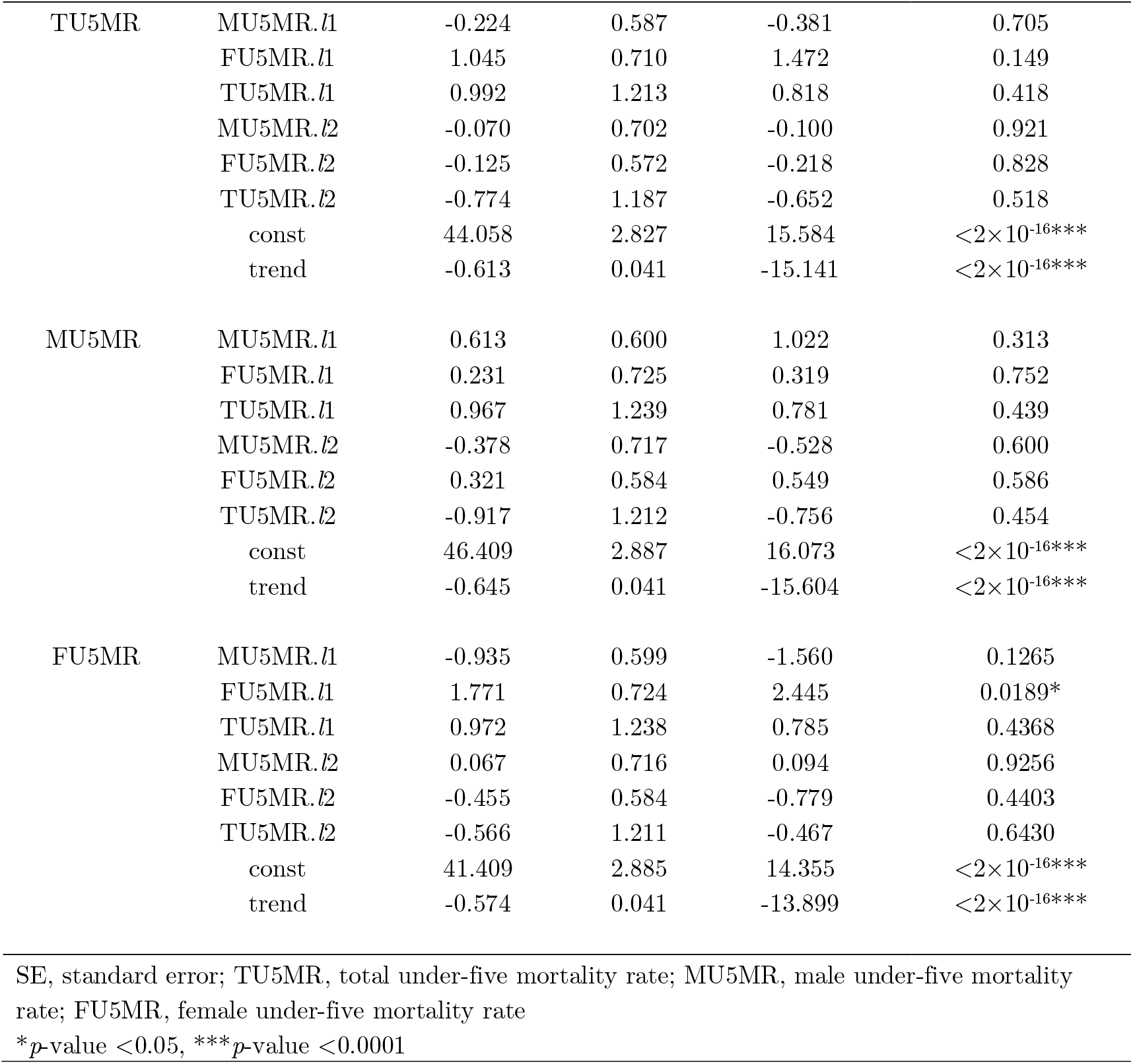
Multivariate estimates obtained using VAR(2) models of TU5MR and its sex composites

#### 3.2.2. VAR model diagnostic tests

The goodness-of-fit of the estimated VAR(2) models constructed in the previous section was further examined using various diagnostic tests, including analyses of serial correlation, heteroskedasticity, residual normal distribution, and structural stability. The serial correlation analysis yielded *χ*^2^ = 171.82 (*p*-value = 0.4623), indicating the non-existence of serial correlations in the estimated VAR models. The heteroskedasticity test also came out with *χ*^2^ = 168.98 (*p*-value = 0.7115), indicating the absence of ARCH effects in the estimated models. The Jarque–Berra test of normality produced *χ*^2^ = 4.71 (*p*-value = 0.5815), which indicated that the resulting study models are characterized by fairly normally distributed residuals. This verdict based on the normality test was also supported by the multivariate tests of skewness and kurtosis which yielded *χ*^2^ = 2.95 (*p*-value = 0.3994) and *χ*^2^ = 1.76 (*p*-value = 0.6236), respectively. Finally, the OLS-CUSUM plots of the empirical fluctuation processes for each under-five mortality rate variable (Fig. 3) was found contained within the 95% confidence bands, indicating the absence of structural breaks in the model residuals. Furthermore, the smaller confidence intervals surrounding the parameter estimate for the entirety of each empiric fluctuation process reflect greater estimation certainty. Generally, the results from the outlined diagnostic examinations gave the indication that the constructed VAR(2) models are adequate for prediction and forecasting.

**Fig. 3.**
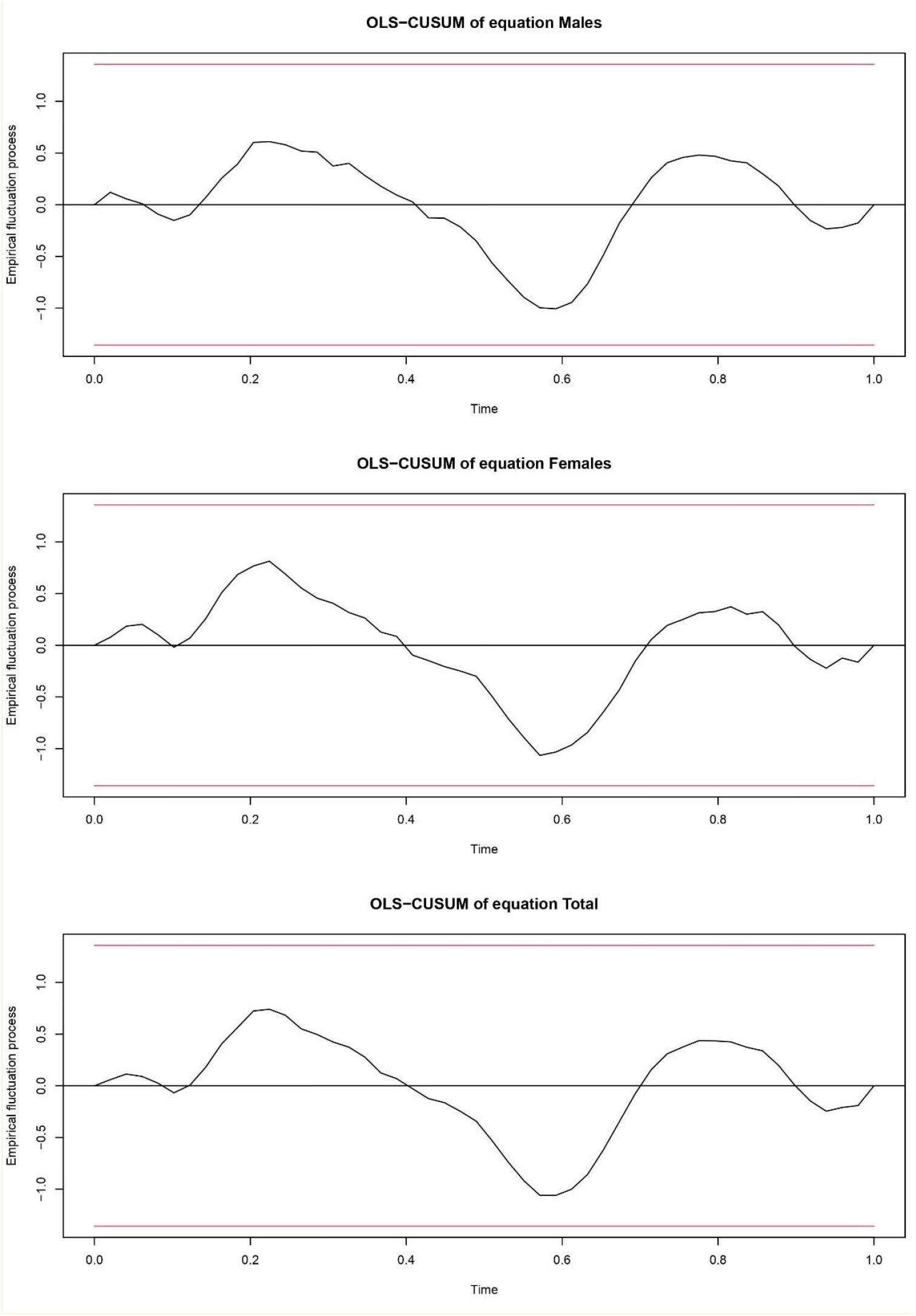
Ordinary least squares (OLS) cumulative sum (CUSUM) plots of parameter estimates of male (top-most), female (middle), and total under-five mortality rates (below) with their 95% level confidence bounds

### 3.4. Instantaneous and Granger-type causality analyses

Table 4 presents the bivariate and trivariate Granger causality analysis results. None of the three mortality indicators Granger caused another in the bivariate analysis (all *p*-values >0.05). However, in the trivariate causality results, all combinations were highly significant (*p*-values <0.0016), except the combination with *H*0: TU5MR ↛ (MU5MR, FU5MR) (*p*-value = 0.8589). This implies that TU5MR did not Granger cause MU5MR and FU5MR, which was intuitive. The counter null hypothesis *H*0: (MU5MR, FU5MR) ↛ TU5MR, however, yielded significant results (*p*-value = 2.2×10^−16^) as expected, implying that MU5MR and FU5MR Granger caused TU5MR. Further, MU5MR Granger caused FU5MR and TU5MR with bidirectionality, as did FU5MR in the Granger causality analysis against MU5MR and TU5MR (Table 4). All trivariate combinations yielded highly significant *p*-values for all null hypotheses with bidirectionality (feedback) (Table 4). These results imply that knowing the future values of any two mortality indicators would help better forecast the other and that future values of any of the three indicators can be forecasted with a smaller forecast error variance if the current, historical, and future values of the other two are incorporated into the model.

**Table 4.**
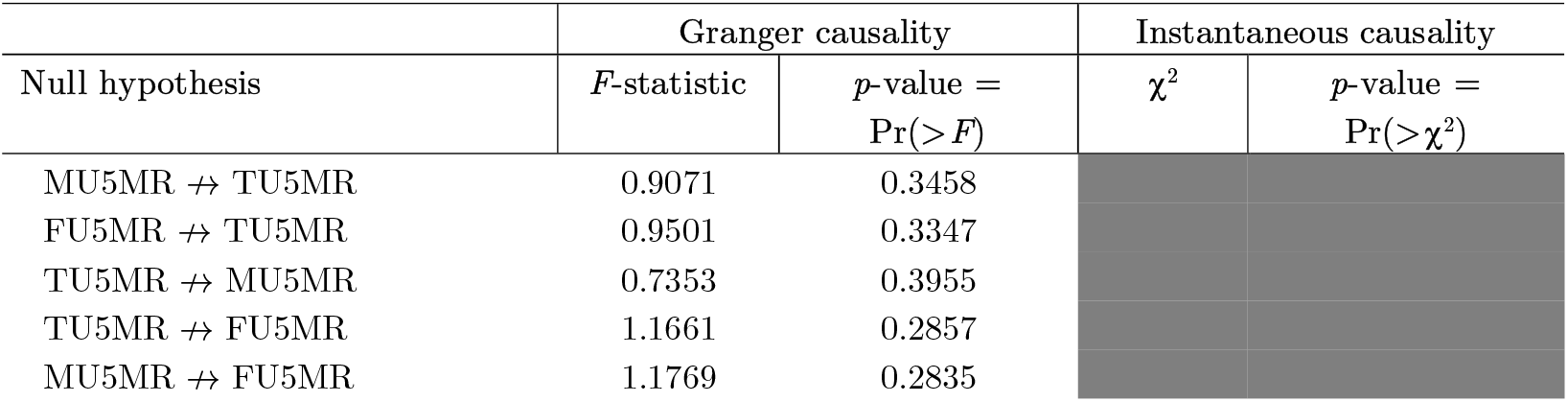
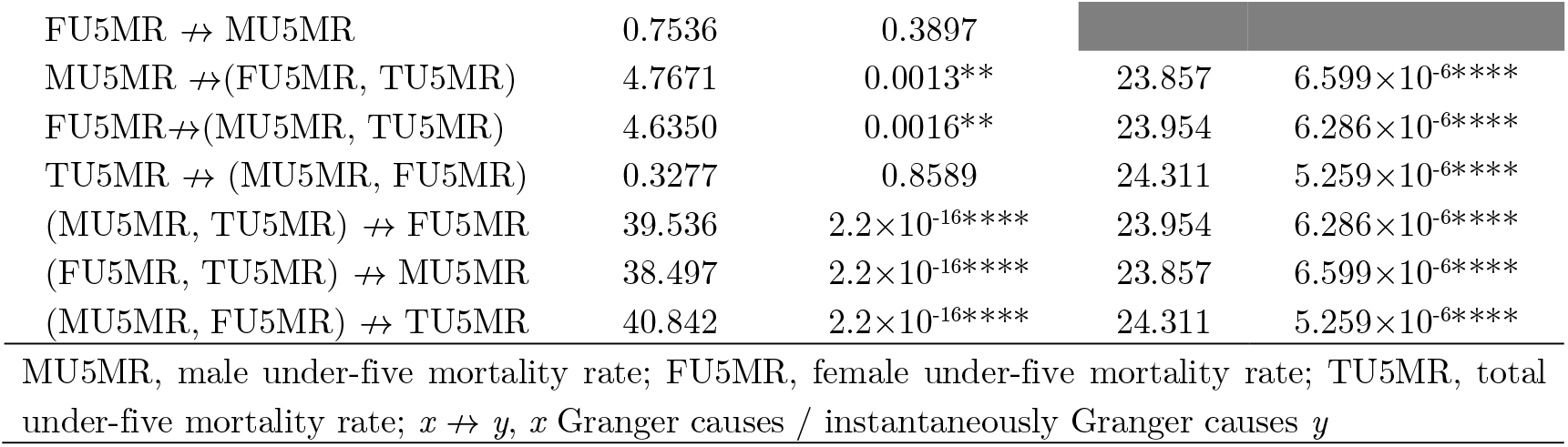
Results of bivariate and trivariate traditional Granger-type and instantaneous causality tests among TU5MR, MU5MR, and FU5MR

### 3.5. Variance decomposition and IRF analyses

Results from the analyses so far have focused on the essential aspects of intrasample tests, which are generally helpful in discerning plausible endogenous Granger-type relations for the period 1970– 2020. However, these results cannot be applied to deducing the extent of exogeneity of the three mortality rates beyond the period of study. To confirm the relative strengths of Granger-type causality, we considered the two approaches, FEVD and IRF analyses, as presented in Section 2.2.3. The FEVD estimated the proportion of each indicator’s forecast error variance that would result from a shock to another within the model while IRF reveals the shocks in the U5MRs. Further, FEVD would indicate the proportion of variation in the forecast error for each mortality rate that could be explained by its own innovations and those of the other rates based on orthogonalized impulse response coefficient matrices. The results of the FEVD analysis are shown in Table 5 whereas those of the IRFs are presented in Fig. 4. The outcomes of both analyses indicate that FU5MR is the most exogenous variable as a high proportion of the shocks shown would be explained by its own innovations in comparison to its contributions to TU5MR and MU5MR (see Table 5). At the completion of 10 years, the forecasted error variance for FU5MR explained its internal innovations by approximately 93% while MU5MR and TU5MR explained their internal innovations by 11.2% and 0.4%, respectively. Further results of FEVD in Fig. 5 show that FU5MR was the most impactful exogenous variable, explaining about 93% of its own internal innovations, unlike MU5MR which only explained approximately 11% at the end of 10 years beyond the historical time series.

**Table 5.** Results of variance decomposition (proportion of forecast variances attributable to internal and external innovations) analyses

| Period | MU5MR | FU5MR | TU5MR |
| --- | --- | --- | --- |
| *Variance decomposition of MU5MR* |  |  |  |
| 1 | 1.0000 | 0.0000 | 0.0000 |
| 2 | 0.9629 | 0.0337 | 0.0033 |
| 3 | 0.8379 | 0.1595 | 0.0025 |
| 4 | 0.6254 | 0.3726 | 0.0019 |
| 5 | 0.4005 | 0.5978 | 0.0016 |
| 6 | 0.2399 | 0.7585 | 0.0015 |
| 7 | 0.1554 | 0.8428 | 0.0017 |
| 8 | 0.1214 | 0.8764 | 0.0021 |
| 9 | 0.1122 | 0.8851 | 0.0026 |
| 10 | 0.1117 | 0.8850 | 0.0031 |
| *Variance decomposition of FU5MR* |  |  |  |
| 1 | 0.7482 | 0.2517 | 0.0000 |
| 2 | 0.6562 | 0.3406 | 0.0030 |
| 3 | 0.5107 | 0.4862 | 0.0029 |
| 4 | 0.3503 | 0.6470 | 0.0025 |
| 5 | 0.2167 | 0.7808 | 0.0024 |
| 6 | 0.1306 | 0.8669 | 0.0023 |
| 7 | 0.0870 | 0.9104 | 0.0025 |
| 8 | 0.0705 | 0.9266 | 0.0028 |
| 9 | 0.0674 | 0.9292 | 0.0032 |
| 10 | 0.0686 | 0.9275 | 0.0037 |
| *Variance decomposition of TU5MR* |  |  |  |
| 1 | 0.9115 | 0.0731 | 0.0153 |
| 2 | 0.8280 | 0.1650 | 0.0070 |
| 3 | 0.6743 | 0.3214 | 0.0043 |
| 4 | 0.4774 | 0.5196 | 0.0030 |
| 5 | 0.2982 | 0.6993 | 0.0024 |
| 6 | 0.1786 | 0.8191 | 0.0022 |
| 7 | 0.1172 | 0.8805 | 0.0023 |
| 8 | 0.0931 | 0.9042 | 0.0026 |
| 9 | 0.0874 | 0.9095 | 0.0030 |
| 10 | 0.0879 | 0.9085 | 0.0035 |

**Fig. 4.**
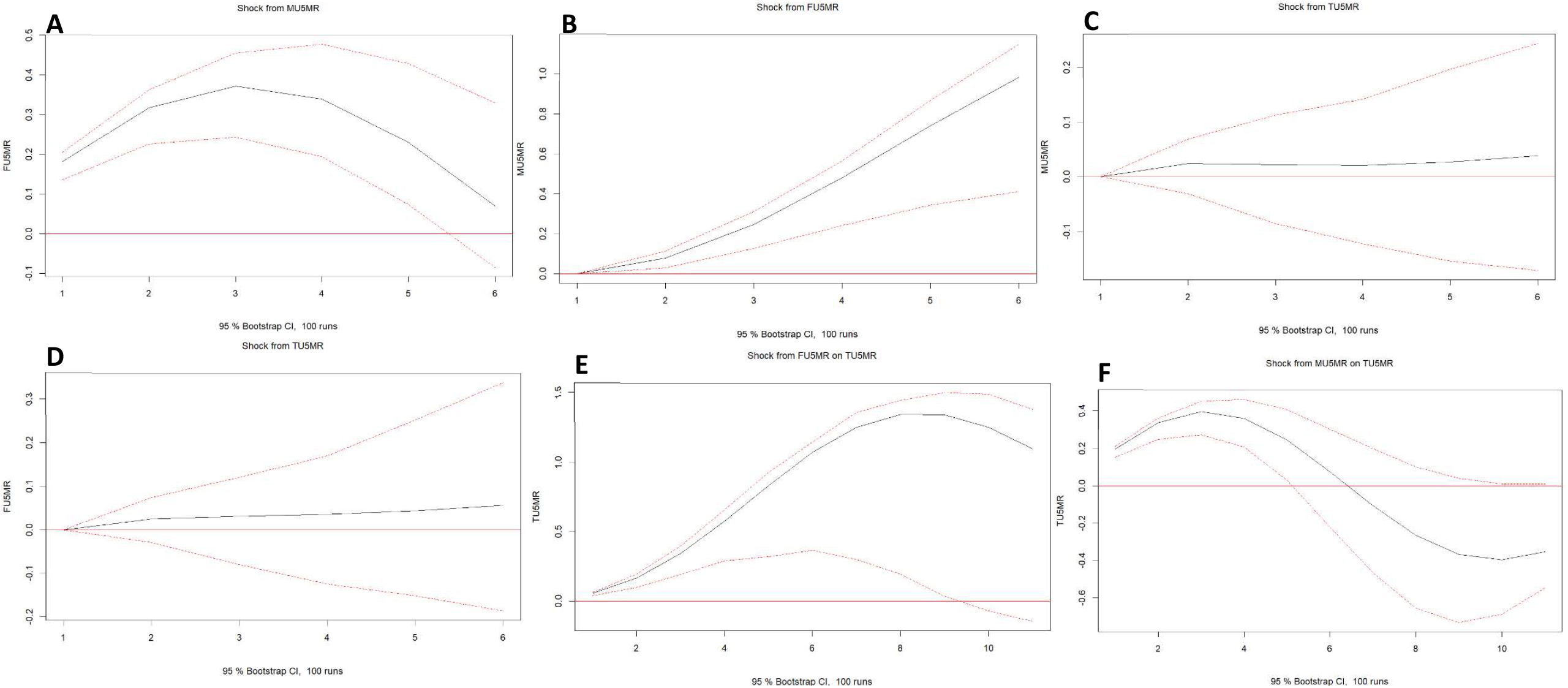
Short-term (5-year) impulse response plots of (A) FU5MR to a 1 standard deviation shock in MU5MR, (B) MU5MR to a 1 standard deviation shock in FU5MR, and (C, D) MU5MR and FU5MR to 1 standard deviation shocks in TU5MR. Also shown are longer-term (10-year) impulse response plots of TU5MR to single standard deviation shocks in (E) FU5MR and (F) MU5MR. MU5MR, male under-five mortality rate; FU5MR, female under-five mortality rate; TU5MR, total under-five mortality rate

**Fig. 5.**
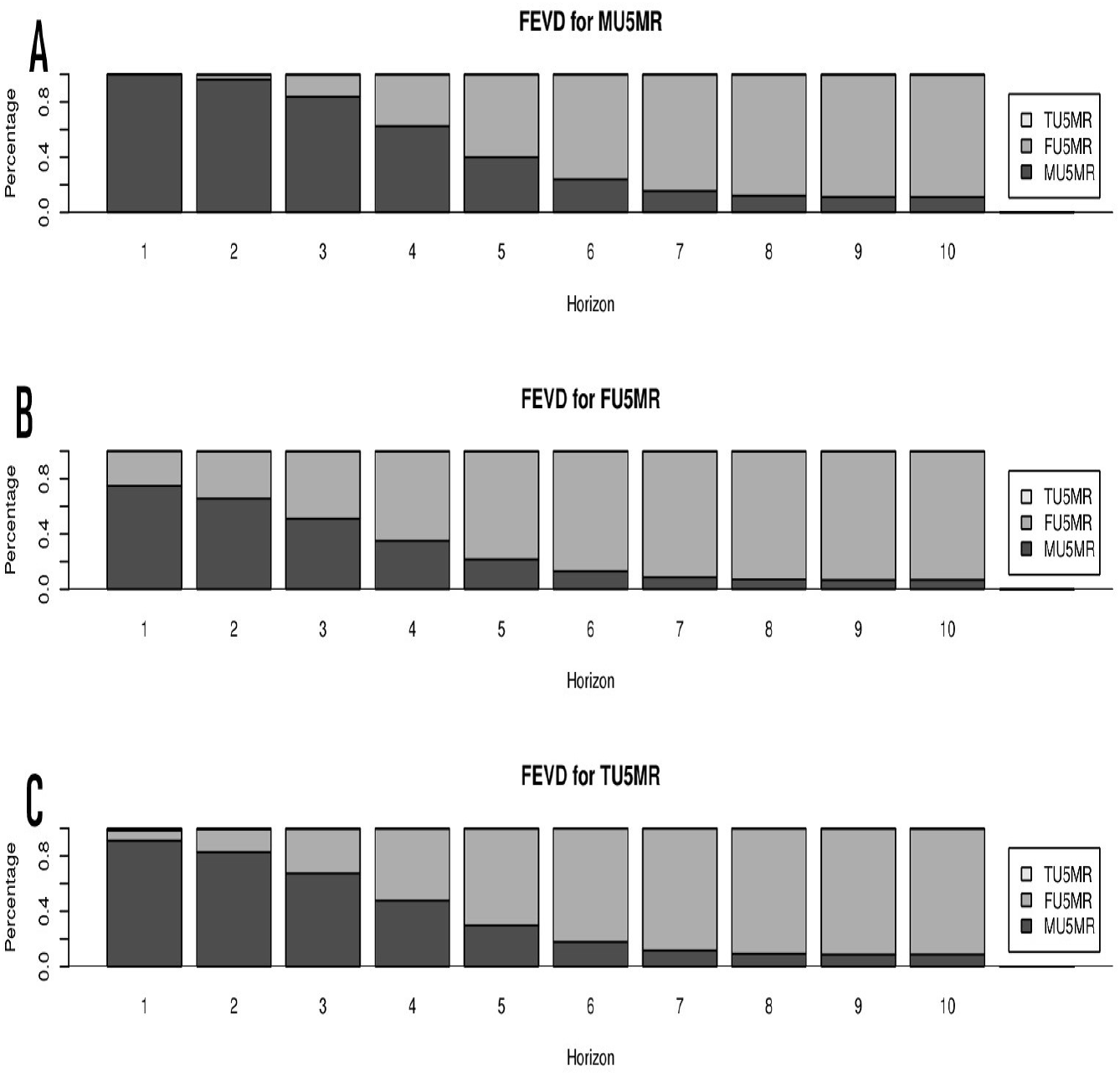
Forecast error variance decomposition (FEVD) plots for (A) male, (B) female, and (C) total U5MRs in Ghana for 10 periods beyond 2020

The short-term (5-year) IRFs (Fig. 4(a)–4(d)) reveal that shocks in the TU5MR had almost no changes on FU5MR and MU5MR, while TU5MR increased sharply when FU5MR was applied as a shocker and increased gradually and declined close to its original values when the MU5MR was applied as a shocker. However, in the long-term (10-year) IRFs (Fig. 4(e) and 4(f)), FU5MR as a shocker resulted in significant increases in TU5MR, while MU5MR as a shocker led to significant decreases in TU5MR in response. Based on the results of FEVD and IRF analyses, it would be reasonable to suggest that focusing on FU5MR would have the most impact on decreasing TU5MR in the Ghanaian context.

### 3.6. VAR forecasting

Table 6 and Fig. 6 show five-year predicted values of TU5MR, MU5MR, and FU5MR from 2021 to 2025 based on the constructed VAR(2) models, which indicate gradual decrease for all the three mortality components. Based on the prediction, from 2021–2025, the TU5MR for Ghana is expected to decrease further by approximately 32%, from 44.71 to 30.38 per 1000 live births (95% CI: 27.84– 32.92), whereas sex-differentiated U5MRs are projected to reach 33.96 per 1000 live births (95% CI: 31.63–36.29) for males and 26.64 per 1000 live births (95% CI: 23.85–29.43) for females.

**Table 6.** Forecasted 5-year total, male, and female U5MRs for Ghana (2021–2025) MU5MR, male under-five mortality rate; FU5MR, female under-five mortality rate; TU5MR, total under-five mortality rate; CI, confidence interval

| Year | MU5MR |  | FU5MR |  | TU5MR |  |
| --- | --- | --- | --- | --- | --- | --- |
|  | Forecast | 95% CI | Forecast | 95% CI | Forecast | 95% CI |
| **2021** | 47.03 | 46.62–47.45 | 38.10 | 37.68–38.51 | 42.66 | 42.26–43.07 |
| **2022** | 44.35 | 43.52–45.19 | 35.77 | 34.89–36.66 | 40.16 | 39.31–41.00 |
| **2023** | 41.19 | 39.92–42.46 | 33.01 | 31.58–34.44 | 37.19 | 35.86–38.52 |
| **2024** | 37.66 | 35.91–39.41 | 29.91 | 27.85–31.97 | 33.87 | 31.99–35.76 |
| **2025** | 33.96 | 31.63–36.29 | 26.64 | 23.85–29.43 | 30.38 | 27.84–32.92 |

**Fig. 6.**
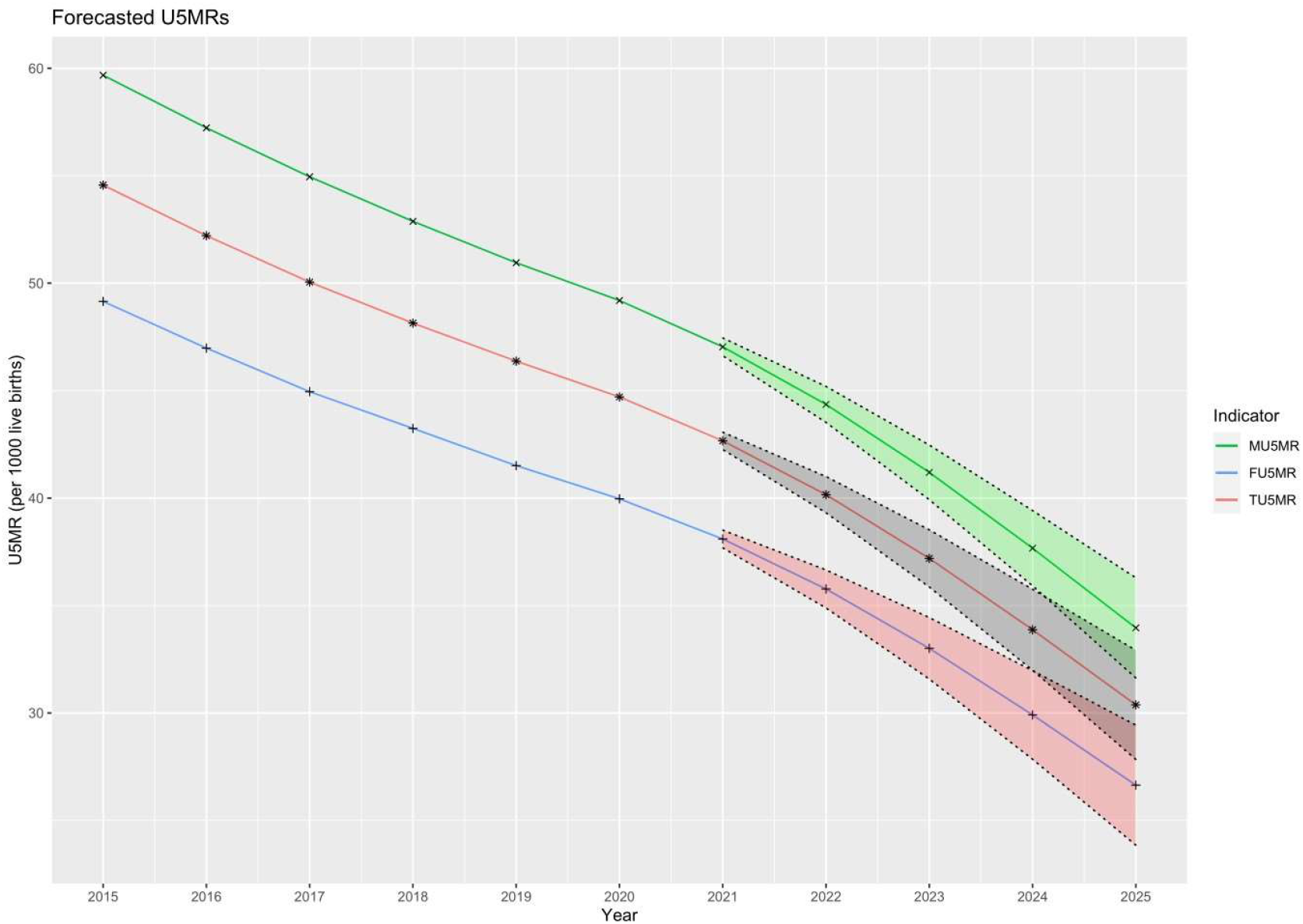
Zoomed-in fan plots showing observed and 5-year VAR(2) forecasts of total, male, and female under-five mortality rates in Ghana (2021–2025) with their respective 95% confidence bands

## 4. Discussion

There has been an urgent need to derive better forms of evidence for resource-limited settings to guide public health systems planning and resource allocation. However, there remains a paucity of studies aimed at obtaining accurate forecasts of child mortality rates in such settings. More specifically, no known study has yet forecasted sex variations in childhood mortality rates or modeled such differences in Ghana. The study was conducted in response to calls in recent times from international monitoring organizations to disaggregate U5MRs by sex [26,27]. We explored a VAR model for analyzing sex disparities in and forecasting U5MR based on 1970–2025 data obtained from the UNICEF Data Warehouse on Ghana. The model produces estimates of U5MR at the national level while capturing sex variations and provides a reproducible approach to projecting U5MR based on the three types of time series as a single vector that is data-driven and modelbased. Validation exercises were also suggestive of considerably adequate calibration and predictive performance.

### 4.1. Study findings

Considering the existing rates of decline, Ghana is likely to achieve the childhood survival goal (SDG 3.2.2) for U5MR of ≤ 25 deaths per 1000 live births before 2030 [28,29], hopefully between 2026 and 2028. This is contrary to the forecasts of Mejía-Guevara et al. [30] and Alhassan et al. [31] who predicted that Ghana would achieve the SDG for U5MR between 2030 and 2050. With regard to sex, the FU5MR may likely reach SDG 3.2.2 between 2025 and 2026, while the MU5MR will likely reach SDG 3.2.2 between 2027 and 2028. Even considering the upper limits of the 95% CIs, the TU5MR will likely reach approximately 25 per 1000 live births between 2027–2028; MU5MR and FU5MR will decline to this level between 2029–2030 and 2026–2027, respectively. Interestingly, our estimated annual rates of change based on historical data between 1970 and 2020 were not significantly different from those estimated by Alhassan et al. [31] based on historical data from 1988 and 2017 (−3.7% vs. -2.99%; 95% CI: -7.5 to 8.92 for FU5MR and -3.1% vs. -3.2%; 95% CI: -7.96 to 7.76). In our estimates, there was no difference between both sexes with respect to the annual decline in U5MR between 1970 and 2020 (*p*-value = 0.9464) and the time series showed significant levels of cointegration.

A gradually declining trend was observed for both male and female children, with MU5MR being consistently higher than FU5MR for both historical and forecasted periods. Despite a mean sex ratio of U5MR of 1.16, the overall sex variations appeared to be relatively stable in historical and forecasted data, unlike countries with similar economic circumstances like Nigeria whose FU5MR has been projected to increase gradually between 2025 and 2030 [32]. Thus, unlike Nigeria, Ghana has shown a relatively well-preserved female survival rate, at least, in terms of U5MR, implying that the biological female advantage has been conserved over the years. Our model also showed that this situation will likely continue in the next 5–10 years, although the gap is gradually collapsing (Fig. 6). It is worth noting, however, that a MU5MR to FU5MR ratio in excess of unity is not necessarily sufficient grounds for concluding that girls do not experience inequity [33]. Girls could show lower U5MRs that are consistently below those of boys while experiencing excess deaths beyond those expected given their genetic and biological survivorship advantage [33–35].

The error variance decompositions revealed the individual effects of MU5MR and FU5MR on TU5MR, with both explaining half of future fluctuations in TU5MR at 4 years. In the longer term (at 10 years), MU5MR and FU5MR explained approximately 9% and 91%, respectively, of future fluctuations in TU5MR. These results confirmed those of the IRF analyses, implying that while focusing on both male and female U5MR could be useful for reducing TU5MR in the short term, child mortality policies that target FU5MR will prove more useful for reducing overall TU5MR in Ghana in the long run.

### 4.2. Strengths and limitations

The present study, to the best of our knowledge, is the first to use a VAR model to forecast TU5MR and its sex differentials (MU5MR and FU5MR) for Ghana. As a strength, the VAR model uses lagged elements of endogenic variables at time *t*; thereby eliminating the simultaneity problem and making the OLS method applicable for estimation [36]. Also, as Verbeek [37] put it, a major advantage of the VAR modeling approach is that it yields a parsimonious model and more precise forecasts because the variables are modeled at the same time, and lagged components render a more informative model. Again, our model was based on annual data, making it possible to capture better trends compared to previous studies [30,31] that attempted to capture these trends using evidence from demographic health surveys whose data are collected at wider intervals.

Notwithstanding, the findings of our study should be construed taking into consideration certain methodological limitations. Despite its elegance in analyzing the short-lag effects of multivariate time series, the VAR model for predicting U5MRs is subject to some limitations as any other correlational analysis. Even though impulse response modeling assesses the direction of impact of each variable on the others, poor model selection threatens the interpretation of our findings. There may exist more variables and measurements that may better inform our conclusions. Further, in line with the saying that correlation does not equal causation, the strength of the VAR model generated herein lies in its ability to establish the correlation between any two of the U5MRs investigated. Beyond this, it yields spurious correlations, as is typical of any such regression model. Despite the aforementioned limitations, the present study provides a nationally representative exposition of TU5MR and sex variations in this important indicator in Ghana and the properties identified herein are generalizable to predictions of short- and long-term U5MRs in Ghana.

## 5. Conclusions

The present study attempts to utilize VAR to model sex differentials in U5MR in Ghana based on data from the UNICEF Data Warehouse from 1970–2020 [14]. It adds to the growing body of studies showing applications of multivariate statistical tools to population health for the effective design, planning, and utilization of health resources in curbing the deaths of children in resource-limited settings like Ghana. In addition to a gradually declining U5MR, overall trends among the sexes were observed to be well maintained in the historical and forecasted data. Further, contrary to the previously published evidence and based on the predictive VAR model established here, Ghana may meet SDG 3.2.2 before 2030 and maintain appreciable declines in U5MR beyond the SDG era if policies and interventions aimed at reducing childhood mortality rates and gender inequities are sustained or scaled up. While the mechanisms underlying sex discriminative practices are complex and range from male/female cultural preference and intentional negligence on the part of parents and caretakers to biases in health resource allocation, these can hardly be investigated entirely based on quantitative modeling. We recommend that further research is necessary to document inequitable allocations of health resources and discriminative treatments between male and female children in Ghana.

## Data Availability

All data produced in the present study are available upon reasonable request to the authors

